# NEURAL CORRELATES OF INFANT SELECTIVE MOTOR CONTROL EMERGENCE WITHIN 5 MONTHS OF LIFE: IMPLICATIONS FOR CEREBRAL PALSY

**DOI:** 10.1101/2025.06.16.25329721

**Authors:** Alexander Drobyshevsky, Vasiliy Yarnykh, Andrea C. Pardo, Raye Ann deRegnier, Colleen Peyton

## Abstract

**Background:** Selective motor control (SMC)—the ability to isolate joint movements—is a critical component of early motor development and a strong predictor of later motor outcomes in individuals with cerebral palsy (CP). While SMC is thought to emerge as descending control from the cortex increases, the neural mechanisms supporting this process in early infancy are not well defined. This study utilizes multimodal MRI to investigate the relationship between microstructural changes in the corticospinal tract (CST) and the development of SMC in preterm infants during the first five postnatal months.

**Methods:** Fifteen preterm infants (<32 weeks GA, <1500g) underwent MRI between 3– 21 weeks corrected age, including macromolecular proton fraction (MPF) mapping to quantify myelination, diffusion tensor imaging (DTI) to assess fractional anisotropy (FA), and g-ratio estimation. SMC was measured using BabyOSCAR, a clinical tool that captures an infant’s capacity to perform isolated joint movements. Regions of interest included the posterior limb of the internal capsule (PLIC; CST) and the corpus callosum. One infant with spastic CP was included for exploratory comparison.

**Results:** In infants with typical development, SMC scores increased with age and were strongly correlated with MPF in the PLIC (R² = 0.81), suggesting close alignment between CST myelination and emerging motor control. MPF in the CST increased rapidly (from 24% to 80% of adult values), outpacing both FA and corpus callosum. In the infant with CP, ipsilesional CST myelination and contralesional SMC scores were substantially lower than expected for their age. Elevated MPF values in the rubrospinal tract suggested compensatory reorganization. In contrast, the infant with early motor delay demonstrated typical MPF, FA, and SMC scores and achieved independent walking by age two, indicating a transient delay rather than evolving CP.

**Conclusion:** Rapid CST myelination during early infancy closely parallels the development of selective motor control. Measures of MPF provide a sensitive structural correlate of this emerging capacity and may help identify early disruptions in motor system development. These findings provide a neurobiological framework for the early detection of motor impairment and more precise timing of intervention in infants with or at an increased chance of CP.

## INTRODUCTION

Cerebral palsy (CP), the most common cause of lifelong motor disability in childhood, frequently results from early brain injury that disrupts the developing motor system^1^. A primary early consequence of such injury, particularly affecting the corticospinal tract (CST), is impaired selective motor control (SMC)^2^—the ability to isolate joint movements crucial for both gross^3^ and fine motor functions^4^. While SMC is a strong predictor of long-term functional outcomes in CP^5^, the developmental trajectory of the increasing descending cortical regulation that underlies SMC remains poorly understood. This lack of understanding represents a significant obstacle to developing timely and targeted interventions.

The early postnatal period is a critical window of neurodevelopment, characterized by rapid and dynamic changes in brain structure and function^6^. During this time, SMC begins to emerge in preterm infants^7^, supporting the development of skilled and dexterous motion. Evidence from animal models suggests that this emerging capacity depends heavily on the maturation of descending cortical pathways, particularly input from sensorimotor cortex to spinal motor circuits ^8, 9^. Despite its importance, the neural substrates underlying this developmental process remain poorly characterized in humans, primarily due to the limited availability of longitudinal neuroimaging studies in early infancy. Identifying the structural correlates of emerging cortical motor control is critical for advancing our understanding of both typical motor development and the early manifestations of cerebral palsy (CP). Magnetic resonance imaging (MRI), particularly diffusion tensor imaging (DTI) and myelin-sensitive techniques, provide a non-invasive means to quantify microstructural and organizational changes within the developing brain that may support the acquisition of motor control.

Prior research has emphasized the importance of white matter maturation, particularly CST development, in motor skill acquisition ^10, 11^. Prior DTI studies have demonstrated a close association between the development of major white matter tracts, including the CST, and the progression of motor abilities ^11^. For instance, increases in fractional anisotropy (FA) and decreases in mean diffusivity (MD) within the CST have been linked to improvements in motor function, reflecting enhanced axonal organization and myelination, which are crucial for efficient transmission of cortical signals ^12, 13^. Furthermore, myelin mapping studies have revealed predictable trajectories of myelination in sensorimotor regions and within the CST during the first year of life, with remarkably rapid changes observed between 1 and 5 months^14^. These findings strongly suggest that the development of cortical control over movement is tightly linked to the structural maturation of key white matter pathways, most notably the CST.

However, a critical gap persists in our understanding of how specific microstructural changes, such as myelination and white matter tract development within the CST, contribute to the increasing capacity of cortical regions to regulate spinal motor circuits and drive the emergence of SMC during the first six months of life. This study investigates the relationship between DTI and myelin mapping techniques and the emergence of SMC in preterm infants during early postnatal development. To measure SMC, we used BabyOSCAR^15^, a clinically validated observational tool^16^ that quantifies joint isolation in infants and predicts CP diagnosis and distribution by age two^5^. By investigating the relationship between white matter microstructure, particularly within the CST, and the emergence of SMC, which reflects increasing cortical control, this research aims to clarify the neural mechanisms underlying early motor development and identify potential biomarkers for detecting infants with probable CP or other motor disorders. The insights gained may also inform the development of early intervention strategies to support optimal motor development in medically complex infant populations.

A specific objective of this study is to evaluate the utility of the macromolecule proton fraction (MPF) for quantitatively assessing cerebral myelination, particularly within the CST, during early postnatal development. Estimation of the myelin water fraction using multicomponent driven equilibrium steady-state observation of T1 and T2^14, 17^ can be achieved in clinically relevant times and has been utilized to track neurodevelopment and neurodegeneration. This approach has also been histologically correlated with myelin content ^14^. This method has been demonstrated to be inaccurate and imprecise when intercompartmental exchange is included in the microstructural model ^18^. Unlike relaxometry-based methods, MPF is less susceptible to the confounding effects of iron deposition. This is particularly relevant given that iron accumulation, potentially resulting from conditions such as interventricular hemorrhage, periventricular hemorrhagic infarctions, and kernicterus in premature infants, is associated with white matter injury, CST damage, and adverse sensorimotor outcomes ^19^.

Simultaneous myelin-sensitive and diffusion tensor imaging enables the in vivo investigation of g-ratio maturation. This crucial microstructural parameter determines the speed and efficiency of signal transmission within white matter tracts. Previous research has observed g-ratio development across early childhood ^20^. We aim to extend this approach to a more granular resolution within the first six postnatal months and investigate its relation to the development of selective motor control in infants born very preterm.

## METHODS

### Participants

Fifteen infants born preterm at <32 weeks’ gestational age and with a birthweight of <1500g were prospectively recruited between June 2022 and March 2023 as a subset of a larger study aimed at assessing the development of SMC. None of the participating infants had congenital malformations or genetic syndromes. The infants underwent MRI under a research protocol approved by a local Institutional Review Board (IRB) between 3- and 21-weeks’ corrected age. Four participants had two serial examinations, resulting in a total of 20 MRI studies. Between 18 and 24 months corrected age, infants received standardized developmental testing as part of their standard of care, with the Bayley Scales of Infant and Toddler Development, 4^th^ edition^21^.

### Selective motor control assessment

The infants were scored every 2-4 weeks from enrollment to 5 months corrected age using video recordings and the Baby Observational Selective Control Appraisal (BabyOSCAR) tool^15^, which assesses joint isolation and the presence of synergies and mirror movements. Two trained raters completed all scoring. The infants were filmed supine during active movement while in a calm and alert state. The scoring is performed on each joint from 1 minute of the recording, and the total combined score ranges from 0 to a maximum of 32.

### MRI Imaging protocol

Subjects were scanned with the parent present on a Siemens 3T system at the Lurie Children’s Hospital Medical Imaging Research Office. An hour before the scan, infants were fed and gently immobilized in the standard manner: swaddled, with their heads on a custom bean cushion, and wearing standard ear protection. Pulse oximetry was used to monitor heart rate and oxygenation throughout the study. No narcotic or sedative agents were administered to any infant for study purposes. MRI images were obtained using a 24-channel head coil array.

#### Macromolecular Proton Fraction (MPF) mapping

A whole-brain 3D MPF mapping protocol was implemented using a standardized acquisition schema ^22^ utilizing a combination of three 3D spoiled gradient-echo pulse sequences (TR/TE/FA): T1- weighted (16/4.6/18), proton density (16/4.6/3), and magnetization transfer (26/4.6/8). All scans were acquired in the sagittal plane with a voxel size of 1.5 × 1.5 × 1.5 mm³, a field of view of 160 × 160 mm, and single signal averaging. The total duration of the MPF scans was approximately 13 minutes. MPF maps were reconstructed using a biophysical two-pool tissue model ^23^ with the previously developed software in the C++ language (available at https://www.macromolecularmri.org/), which implements a single- point algorithm with a synthetic reference image ^23^. MPF has been extensively validated as a quantitative metric of myelination due to the strong correlations between MPF and myelin content in neural tissues, as reported in several studies ^24^. This rapid technique is clinically feasible and has been implemented on clinical scanners to assess myelination in fetuses ^25^, infants ^26^, and young children ^26^. The accuracy of the method in depicting differential regional developmental trajectories in gray and white matter ^27^ has been validated using quantitative histology and electron microscopy.

Diffusion tensor imaging (DTI) was performed with a Single-shot EPI pulse sequence in axial slice orientation, 1.92x1.9 mm in-plane spatial resolution, 68 slices 1.9 mm thick, FOV 180x140 mm, matrix 94x94, TR/TE = 8000/74 ms, NEX=1, SENSE 2, 64 non-collinear diffusion directions with b values 0 and 1000 sec/mm^2^. The orientation of the slices was parallel to the line connecting the anterior and posterior commissures (AC-PC). The acquisition time was 9 minutes and 21 seconds. An additional scan with b=0 and reversed phase encoding direction was acquired and used for distortion correction using the FSL topup tool. The diffusion tensor imaging data were skull stripped, and fractional anisotropy (FA), mean, and directional diffusivity maps were calculated using the FSL diffusion toolbox.

### Aggregate g-ratio mapping

The aggregate g-ratio, defined as the inner-to-outer diameter of myelinated axons in a given voxel, was estimated from single-shell DTI^28^ as follows: *g*- ratio=√1/(1+MVF/AVF)^29^, where MVF is a myelin volume fraction (MVF) and AVF is axonal volume fraction. MVF maps were derived from MPF using geometric analysis, scaling factor suggested by ^30^. The corresponding AVF map was calculated from the MVF, and the intracellular volume fraction (ICVF) maps as follows: AVF = (1 − MVF) × ICVF. The ICVF map was derived from NODDI-DTI ^31^, a single-shell diffusion approximation of neurite orientation dispersion and density imaging (NODDI), using an in-house MATLAB script.

### Region of interest (ROI) determination

MPF maps were aligned to DTI scans of the same subject using the FSL non- linear registration tool. Using ITK-SNAP software (http://www.itksnap.org/) and a directional color-encoded FA map, regions of interest (ROIs) were placed bilaterally on the posterior (PLIC) and anterior (ALIC) limbs of the internal capsule, corpus callosum, on a slice crossing the thickest portion of the corpus callosum splenium (CCS) (Figure 1). To assess pathways controlling limb movements, the PLIC was divided into four equal segments, and ROIs were placed on the third and fourth segments, which carried descending projections for the arms and legs ^32^. The rubrospinal tract was delineated by placing ROIs on the red nucleus guided by anatomical landmarks ^33, 34^. Averaged FA and MPF values were extracted from the ROIs in white matter tracts. The MPF and FA values were also normalized to the range of changes during development, based on literature values in typically developing fetuses and adults ^25, 35^, and expressed as a percentage of change from the minimum to the maximum value.

**Figure 1.**
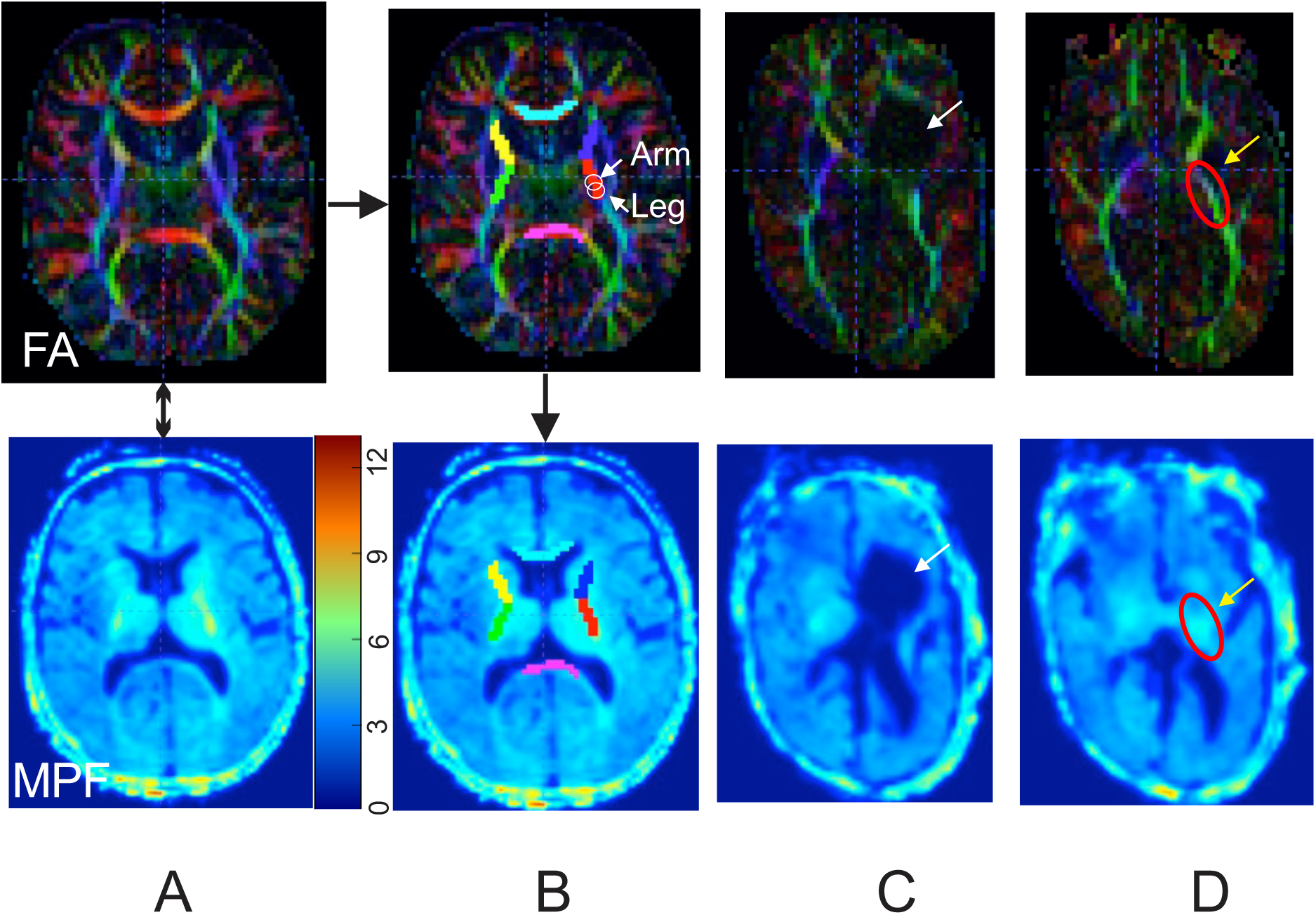
Multimodal evaluation of major white matter tracts in a typically developing infant at 17 weeks CGA (A, B) and in an infant with CP at 9 weeks CGA (C, D). A. FA and MPF maps were spatially co-registered. B. Regions of interest (ROIs) were manually outlined bilaterally on color directionally encoded FA maps (top row) and transferred to the co-registered MPF maps (bottom row). The color bar indicates macromolecule proton fraction mapping in percentage. Portions of PLIC were also delineated for CST portions that predominantly carried motor fibers controlling the arms and legs. C. Color directionally encoded FA maps in an infant with CP with periventricular leukomalacia at 9 weeks. The white arrow on the FA map indicates the location of the lesion and injury to PLIC. ROIs were placed on a slice adjacent to the lesion where the CST tract was visibly preserved in the same infant with CP (D).

## RESULTS

Out of 20 MRI studies performed for this project, several were partially excluded due to excessive motion, poor image quality, or data loss during archival. The resulting 14 DTI scans and 15 MPF maps were used for analysis. The demographic and clinical characteristics are summarized in Table 1. In the infants participating in the study, most had typical motor development (n=13, 87%), as evaluated by the Bayley Scales of Infant and Toddler Development, version 4 ^36^ at ≥18 months corrected age, except for one infant with gross motor delay at 18 months, and one infant with spastic cerebral palsy who had a history of unilateral IVH grade IV, periventricular leukomalacia, and ventriculoperitoneal shunt placement. This subject was later characterized as having a Gross Motor Functional Classification Scale level IV (at 2 years corrected age) with bilateral involvement of the lower extremities and involvement of the right upper extremity. Data from participants with atypical motor development (n=2) were excluded from the calculation of age-dependent regressions for infants with typical motor development and are plotted on graphs separately.

**Table 1.** Demographic and clinical characteristics of the study participants. The data are expressed as median value (interquartile range)

|  |  |
| --- | --- |
| Number of participants | 15 |
| Gestational age (weeks) | 29 (28, 30) |
| Range of gestational age, weeks (min, max) | 24, 31 |
| Birthweight (g) | 1080 (945, 1285) |
| Female | 6 (40) |
| Race |  |
| Asian | 3 (20) |
| Black or African American | 1 (7) |
| White | 9 (60) |
| Unknown | 2 (13) |
| Ethnicity |  |
| Hispanic or Latino | 2 (13) |
| Not Hispanic or Latino | 12 (80) |
| Unknown | 1 (7) |
| Intraventricular Hemorrhage | 2 (13)<sup>a</sup> |
| Bronchopulmonary dysplasia<sup>b</sup> | 10 (67) |
| Treated retinopathy of prematurity<sup>c</sup> | 3 (20) |
| Necrotizing enterocolitis<sup>d</sup> | 0 (0) |

<sup>a</sup>One subject with grade II, one subject with grade IV <sup>b</sup>Required supplemental oxygen ≥35 weeks gestation, <sup>c</sup>Treated with Avastin and/or laser therapy, <sup>d</sup>Treated with drain or laparoscopy

### Selective motor control improves with age, but its emergence differs in individuals with cerebral palsy

Indexes of selective motor control in infants with typical motor development gradually increased within the studied age period, from 1 to 5 months of corrected gestational age (CGA). The BabyOSCAR score was normalized to the maximum value for this age range (7 for legs and 9 for arms) and shown in Figure 2 as a percentage value. The slope of regression for legs was 3.44 % per week, R^2^ = 0.68, and for arms, the slope was 3.70 % per week, R^2^ = 0.75, p-value for both slopes being non-zero was <0.0001. Indexes of SMC for arms and legs in the infant with CP were below 95% confidence intervals of typical development for the corresponding age, except for the score in the ipsilesional left arm. Indexes of SMC for arms and legs in the infant with motor delay were within the range of 95% confidence intervals of typical development.

**Figure 2.**
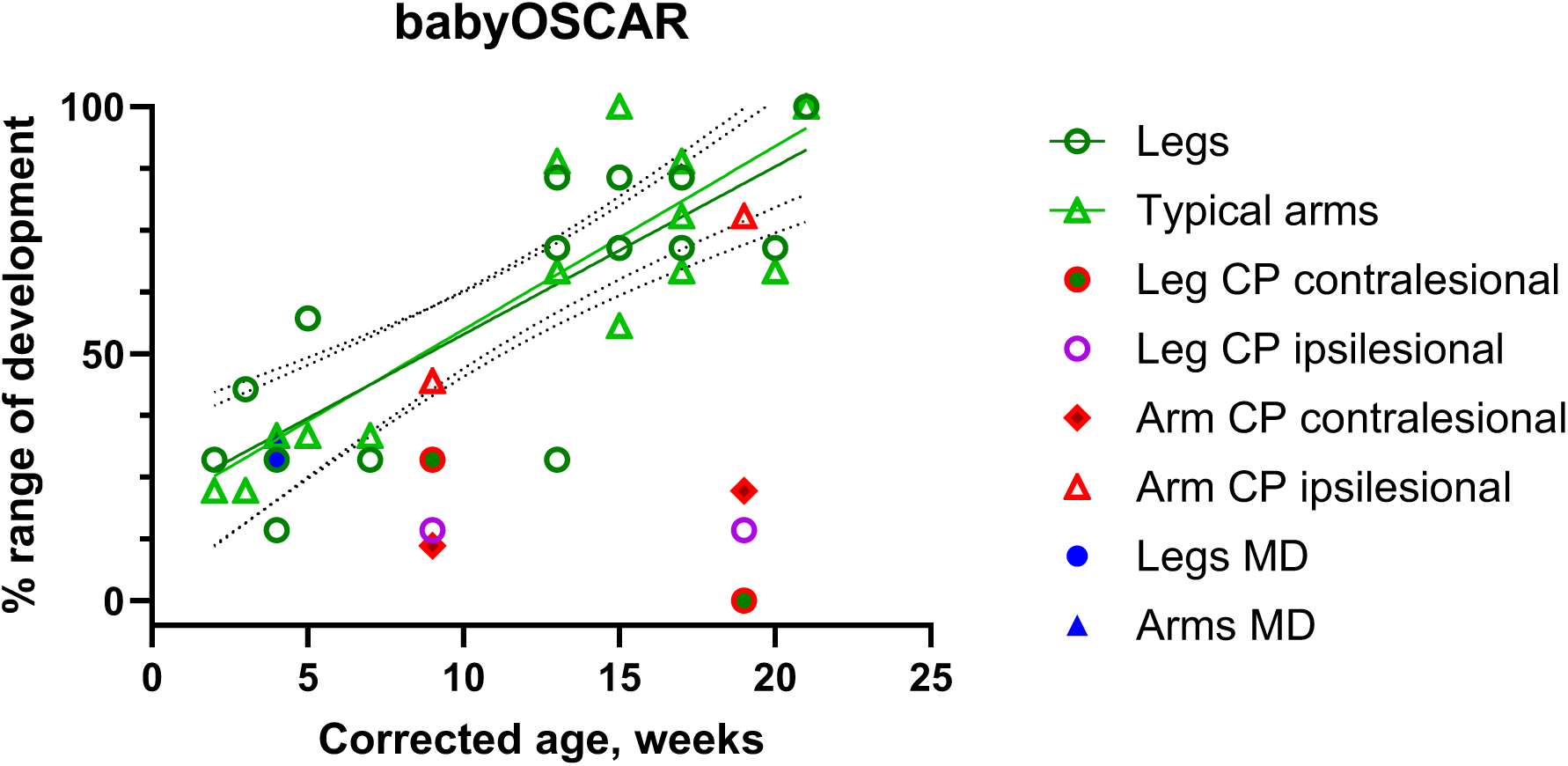
Selective motor control increases in arms and legs in typically developing children and infants with cerebral palsy. The linear regression line and 95% confidence intervals are shown for the arms and legs assessment. Data for the typically developing infants are shown as averages from the left and right sides. Data for the infant with cerebral palsy (CP) are presented separately for the right (contralateral to the lesion) and left (ipsilateral to the lesion)

### Rapid corticospinal tract myelination, detectable with MPF, outpaces callosal development in early infancy

Myelination (measured by MPF) of the PLIC progresses from 24% to 80% of maximum values within the first five postnatal months in infants with typical motor development when normalized to the reported range of changes across the lifespan^37^. In contrast, the corpus callosum, a commissural tract with protracted structural and functional maturation ^38^, shows a more minor MPF change during the same period, from 0.5% to 32% of the maximum adult value. The slope of regression for MPF PLIC was 3.36 % per week, R^2^ = 0.84, and for MPF CC, the slope was 1.37 % per week, R^2^ = 0.56, p-value <0.0001. The slope of regression for MPF in the PLIC was significantly larger than in CC, F_1,32_=19.28, p=0.0001. In the infant with CP, MPF values in the PLIC ipsilateral to the lesion were considerably outside the 95% confidence interval of the regression line. In contrast, the MPF in the contralateral PLIC of the infant with CP fell within the range typically observed in developing infants (Figure 3A).

**Figure 3.**
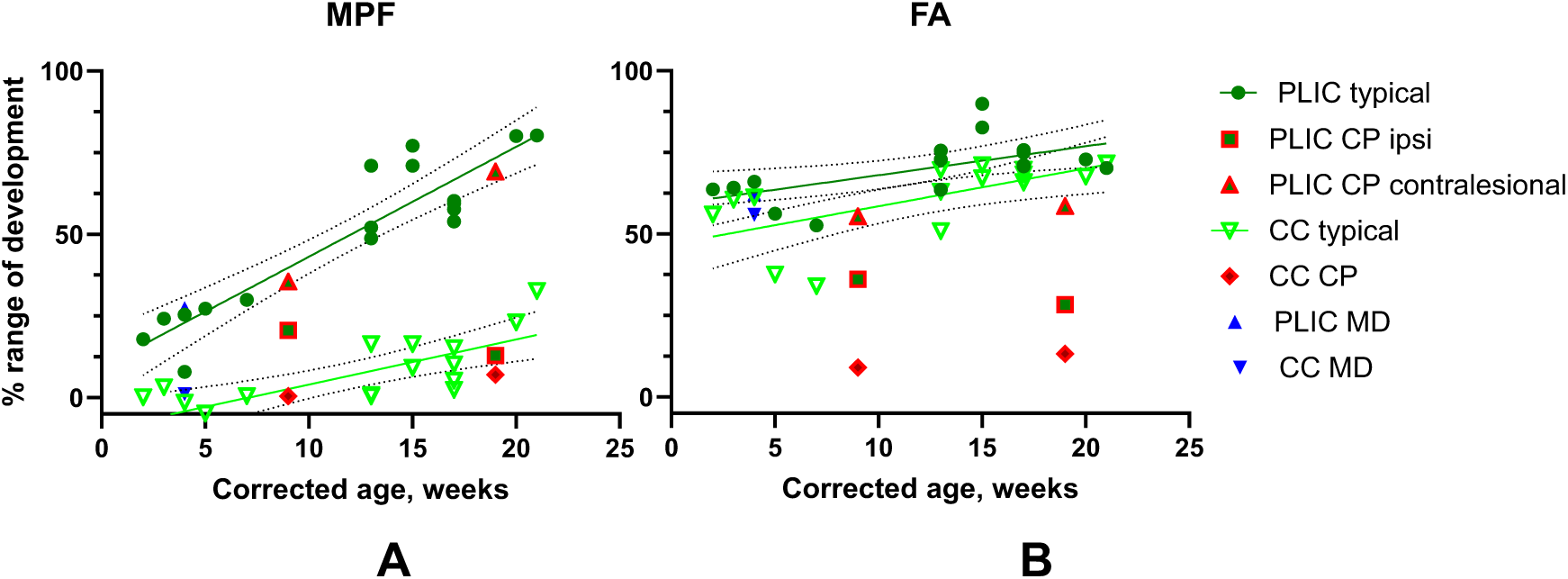
Developmental changes in MPF(A) and FA(B) in the posterior limb of the internal capsule and the corpus callosum in typically developing children and infants with cerebral palsy. The linear regression was calculated for the infants with typical motor development, and 95% confidence intervals are shown. Data for the typically developing infants are shown as averages from the left and right sides. Data for the CP patient are shown separately for the right (contralateral to the lesion) and left (ipsilateral to the lesion) sides, marked with a red border.

In contrast to MPF, the increase in FA in the first 5 months started from a higher value, was of lesser magnitude, and was similar in the PLIC and the corpus callosum, ranging from 52% to 89% for the PLIC and 37% to 71% in the corpus callosum (Figure 3B). The slope of regression for FA in PLIC was 0.88 % per week, R^2^ = 0.35, and for CC, the slope was 1.16 % per week, R^2^ = 0.40, p-value <0.01. The slope of regression for FA in the PLIC was not different from that in CC, F_1,28_=0.305, p=0.58. Like MPF, the PLIC ipsilateral to the lesion was considerably outside the 95% confidence interval of the regression line. At the same time, FA in the contralateral PLIC fell within the range for typically developing infants. MPF in the corpus callosum of the infant with CP was within the range for typically developing infants. The FA in the corpus callosum of the infant with CP was also significantly lower than in typically developing infants. Indexes of FA and MPF in the infant with motor delay were within the range of 95% confidence intervals of typical development.

Taken together, these data indicate that myelination in the corticospinal tract is the process that starts and occurs predominantly within 5 months after term age. At the development of structural organization in white matter, as measured by the FA index, is already well advanced between 1 and 5 months and occurs at a significantly slower rate than MPF. Therefore, myelination in the corticospinal tract, as described by MPF, is positioned as a structural MRI index that may coincide with and indicate the acquisition of functional milestones in motor development, such as the emergence of selective motor control.

### Myelination in the corticospinal tract closely corresponds to the development of selective motor control

Total BabyOSCAR scores (combined score for the limbs), normalized to the maximum value for studied period increased proportionally with age in infants with typical motor development and more closely corresponded to normalized MPF values in the PLIC than to the normalized values of FA (a dashed identity line is shown in Figure 4A). Total BabyOSCAR scores were strongly correlated with PLIC MPF (r=0.91, R^2^=0.81, p-value <0.0001), and moderately correlated with PLIC FA (r=0.69, R^2^=0.47, p-value =0.003). To characterize developmental changes in the infants with CP and motor delay, deviations in behavioral and MRI indexes from values predicted for the corresponding age by linear regression in typically developing infants are plotted in Figure 4B and C. The values of the behavioral and MRI indexes fell in the lower-left quadrant, indicating a delay in functional and structural development of the corticospinal tract. This delay increased with age between 9 and 19 weeks, as indicated by the corrected age values in the serial exams for this patient, which deviated further from zero, where most values for typically developing infants typically concentrate. As expected, the arm contralateral to the PVL lesion in this infant with CP was more affected, as indicated by both BabyOSCAR scores and MPF/FA changes in the corresponding corticospinal tract’s portions.

**Figure 4.**
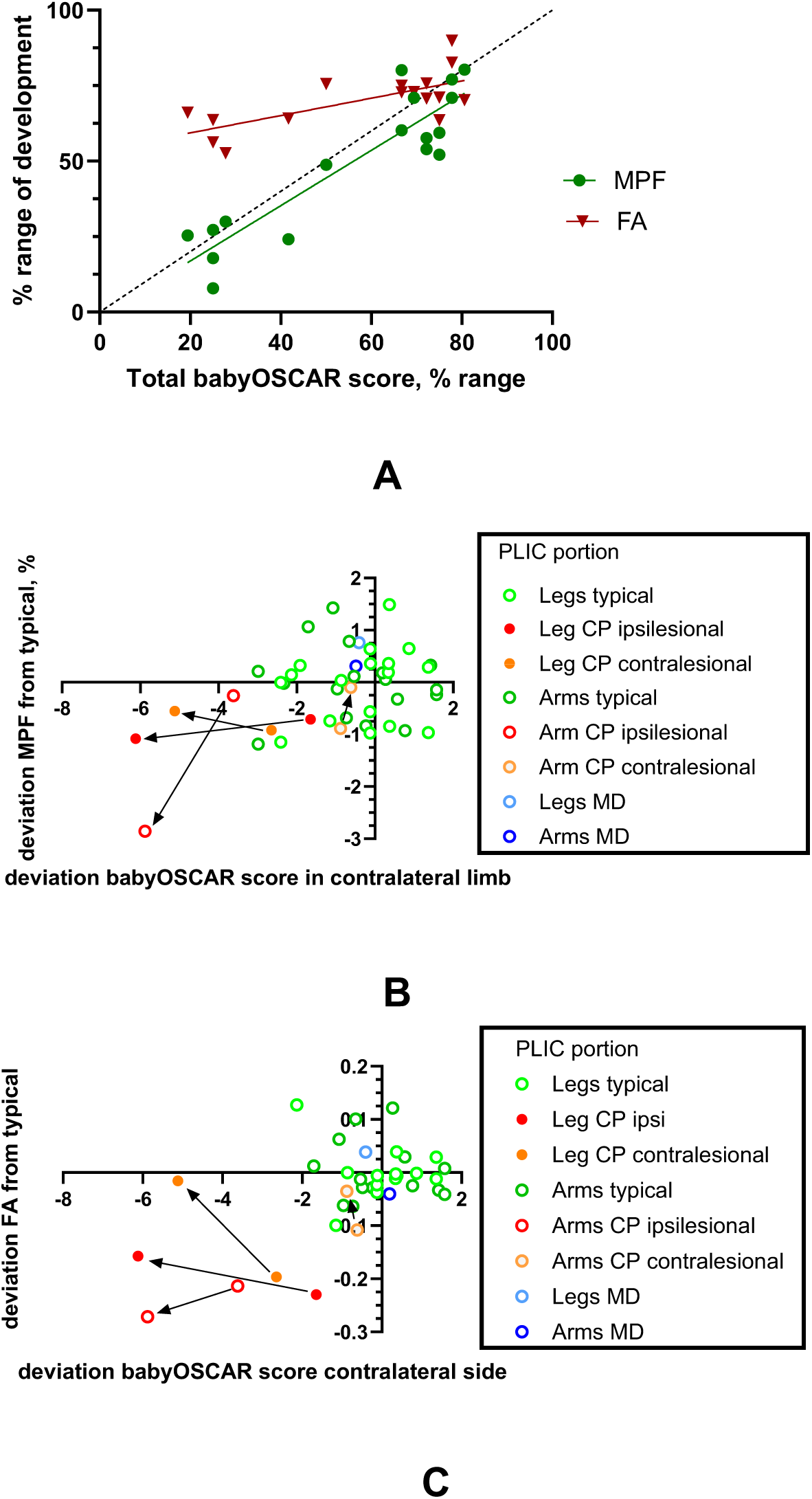
Relationship between selective motor control and MRI indices in the posterior limb of the internal capsule (PLIC). A.MPF and FA indices for the typically developing infants are shown as averages from the left and right sides. The identity line is marked as dashed. Linear regressions were estimated for BabyOSCAR, MPF, and FA vs. age between 3 and 21 weeks of CGA in typically developing infants. Deviations of the actual form from the predicted values in each limb for the corresponding age are plotted for MPF (B) and FA (C) for the typically developing infants, the infant with motor delay (MD) and the infant with cerebral palsy (CP). BabyOSCAR score is plotted for the limb contralateral to internal capsule side on MRI. Arrows indicate the transition between the first and the second MRI study at 9 and 19 weeks in the infant with CP.

### G-ratio development in projection and commissural fibers

The axonal fraction, estimated from NODDI-DTI, demonstrated a slow increase in PLIC between 3 and 21 weeks of corrected age for typically developing infants (Figure 5A). The slope of regression for axonal fraction in PLIC was 0.96E-3 per week, R^2^ = 0.13, not significantly different from zero, and in CC, the slope was 5.8E-3 per week, R^2^ = 0.68, p-value <0.0001. In the infant with CP, the axonal fraction was lower than in typically developing infants of the corresponding age and fell below the 95% confidence interval line. In contrast, the developmental change in the axonal fraction was larger, with a slope difference F1,28=15.2, p=0.0005, suggesting active axonal growth in this age period for commissural tracts. In contrast, axonal growth is nearly complete in projection tracts, such as PLIC. The value of axonal fraction in the corpus callosum was within the range of the typically developing infants.

**Figure 5.**
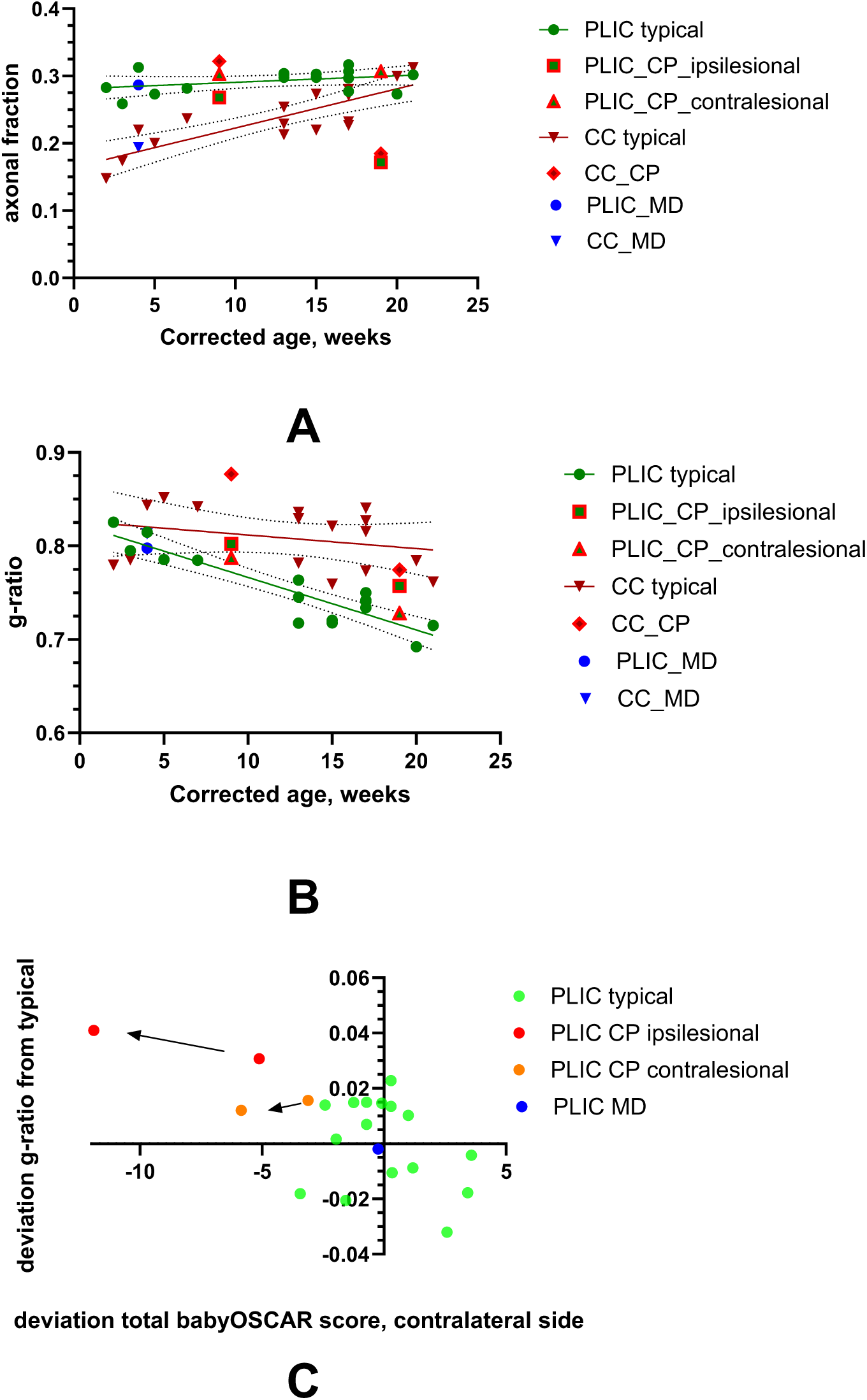
Developmental changes in axonal fraction (A) and g-ratio (B) in the posterior limb of the internal capsule (PLIC) and the corpus callosum (CC) in typically developing infants, the infant with motor delay (MD), and the infant with cerebral palsy (CP). The linear regression line and 95% confidence intervals are shown. Data for the typically developing infants are shown as averages from the left and right sides. Data for the infant with CP are shown separately for the contralesional and ipsilesional, marked with a red border. C. Deviations of the actual g-ratio form from the predicted values using linear regression vs. age between 3 and 21 weeks of CGA for the typically developing infants and the infant with CP. BabyOSCAR score is plotted for the limb contralateral to internal capsule side on MRI. Arrows indicate the transition between the first and the second MRI study at 9 and 19 weeks CGA in the infant with CP.

corpus callosum (Figure 5B), slope difference F_1,28_=7.73, p=0.0096, and almost reached the optimal adult values around 0.7 ^39, 40^. The slope of the regression for g-ratio in PLIC was -5.6E-3 per week, with R2 = 0.82, p-value < 0.0001. In contrast, the slope in CC was -1.45E-3 per week, which was not significantly different from zero, with R^2^ = 0.078. The g-ratio in the ipsilesional PLIC in the infant with CP was above the 95% confidence interval line both at 9 and 19 weeks, while the g-ratio in the contralesional PLIC was within the 95% confidence interval line. The deviation from the estimated values of the g-ratio in the infant with CP from the typically developing infants in the ipsilesional PLIC corresponded to the larger deviation in the BabyOSCAR score, increasing between the 2- and 5-month visit (Figure 5C).

### Rubrospinal myelination increases with age and is elevated in CP

Myelination values associated with MPF increased with age in the rubrospinal tract of typically developing infants between 3 and 21 weeks corrected age (Figure 6A), while FA in the rubrospinal tract demonstrated no apparent change at this period (Figure 6B). Myelination in the rubrospinal tract of the infant with CP had a higher value than that of those in typically developing infants, which may reflect greater reliance on the bulbospinal pathways with the injury to the corticospinal tract.

**Figure 6.**
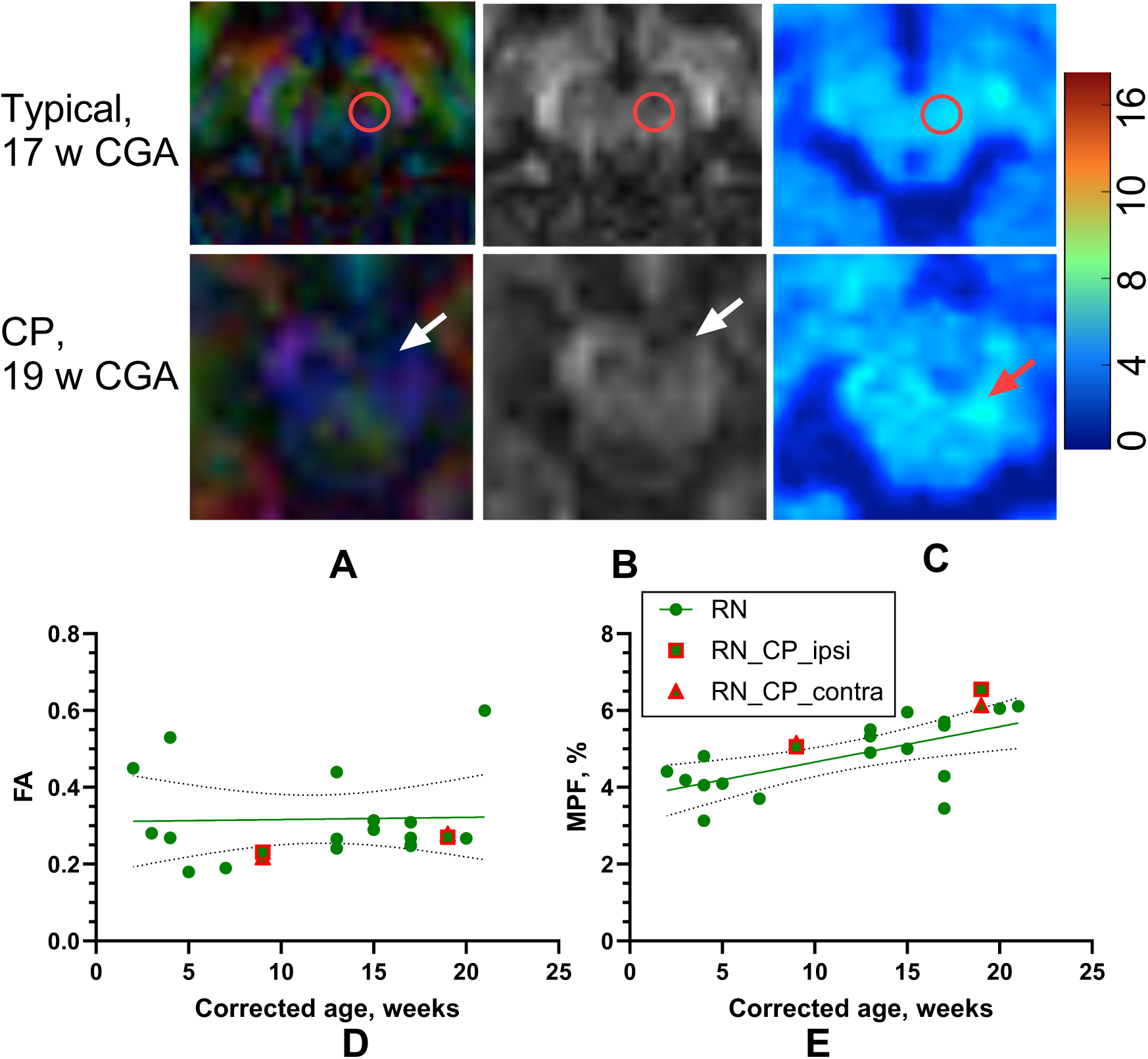
Rubrospinal tract between 2 and 21 CGA. The rubrospinal tract was delineated by placing ROIs on the red nucleus in the mesencephalon and adjacent slices using a color- encoded FA map (A, red circle) and transferred to the FA map (B) and MPF map (C). The color bar indicates macromolecule proton fraction mapping in percentage. The top row shows an infant with typical motor development at 17 weeks corrected gestational age (CGA), and the second row shows an infant with cerebral palsy (CP) at 19 weeks CGA. Note decreased anisotropy in ipsilesional cerebral peduncle (white arrow) and increased MPF in rubrospinal tract (red arrow). A, B. - Developmental changes in MPF(A) and FA(B) in typically developing infants and in the infant with cerebral palsy. The linear regression line and 95% confidence intervals are shown. Data for the typically developing infants are shown as averages from the left and right sides. Data for the infant with cerebral palsy (CP) are presented separately for the right (contralateral to the lesion) and left (ipsilateral to the lesion) sides, marked with a red border.

### Myelination gradually increases between 3 and 21 weeks in the cerebral cortex

Myelination gradually increased with age between 1 and 5 months in all areas of the cerebral cortex, as shown for the precentral and anterior cingulate regions in Figure 7. There were no apparent differences in MPF changes between cortical areas related to motor activity and other cortical areas indicative of emerging selective motor control. Notably, cortical myelination contralateral to the lesion in CP was increased, as shown on MPF maps in Figure 5B and C for the CP infant at two serial studies.

**Figure 7.**
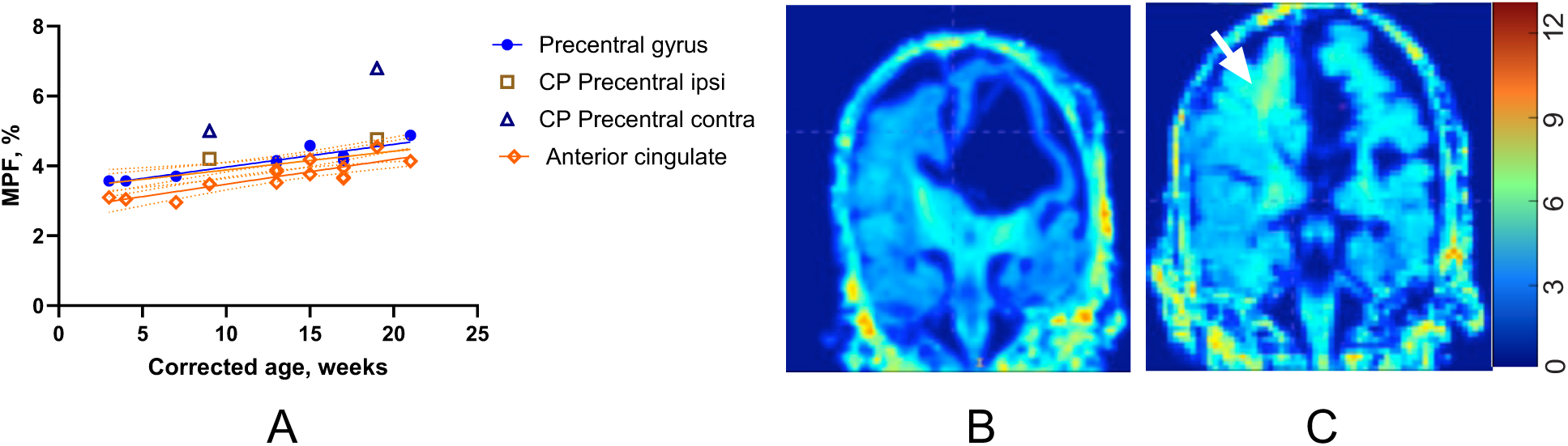
Cortical myelination between 3 and 21 weeks CGA. A. MPF changes in motor and nonmotor cortex with age in the typically developing infants and infants with CP ipsi- and contralateral to the lesion. MPF maps on studies at 9 (B) and 19 (C) weeks. The arrow indicates increased MPF in the perirolandic area.

## DISCUSSION

This study utilized multimodal MRI techniques to characterize the intricate relationship between early brain development and the emergence of SMC in typically developing infants, a child with motor delay, as well as a single case of an infant with spastic CP following intraventricular hemorrhage and periventricular leukomalacia. Our findings highlight the differential developmental trajectories of various white matter indices and their distinct relationships with isolated joint control with a particular emphasis on the role of myelination in the CST.

### Differential Maturation of White Matter Tracts

Our data reveal that myelination, as quantified by MPF, undergoes a significant and accelerated increase within the CST, as measured within the PLIC during the first five months after term age. This rapid maturation contrasts sharply with the corpus callosum, a commissural tract known for its protracted structural and functional development^38^, which showed a much smaller MPF change (0.5% to 32% of maximum adult value) during the same period. This suggests that while the corpus callosum is in its early stages of myelination, the CST is undergoing a critical and rapid phase of myelination during early infancy.

In contrast to MPF values, FA, which reflects the structural organization and coherence of white matter, presents a different developmental profile. FA started from a higher baseline, exhibited a smaller magnitude of increase, and showed similar developmental patterns in both the PLIC (52-89%) and the corpus callosum (37-71%) within the first six months (Figure 3B). This indicates that while structural organization (FA) is already relatively advanced early on, myelination (MPF) is the more dynamic and rapidly progressing process within this specific developmental window. This pattern suggests that myelination, as quantified by MPF, may be a more functionally relevant marker of activity-dependent maturation within the CST during early infancy than FA, which reflects broader aspects of white matter organization but is less directly tied to use-dependent change. Myelination measures of MPF offer a means to assess the development and functional activation of the corticospinal system, as oligodendrocytes preferentially myelinate electrically active axons^41^. This interpretation is consistent with prior research demonstrating the sensitivity of MPF to regionally specific developmental trajectories in both gray and white matter.^42^

### Myelination and Selective Motor Control

A key finding of this study is the high correlation between the rapid myelination of the CST, as measured by MPF in the PLIC, and the emergence of selective motor control. Normalized Total BabyOSCAR scores (combined for limbs) increased proportionally and aligned more closely with normalized MPF values in the PLIC than with FA (Figure 4A). This supports our hypothesis that the development of selective motor control is intrinsically linked to the increasing descending functional drive from the sensorimotor cortex, whose maturing projections form the CST. Efficient, fast conduction along these myelinated fibers is essential for the precise timing and control required to achieve isolated joint movement. Given that oligodendrocytes preferentially myelinate electrically active axons^41^, MPF may also serve as a proxy for circuit engagement, offering insight into both structural development and functional use. These data support MPF as a promising structural MRI index that aligns with the emergence of individuated joint movements driven by corticospinal circuit maturation.

### Insights from the Cerebral Palsy Case Study

The single case of the infant with spastic CP provided valuable preliminary insights into atypical development. This infant, with a history of IVH grade IV and PVL, showed significant differences in both behavioral (BabyOSCAR scores) and structural/functional MRI indices (MPF, FA, g-ratio) in the PLIC ipsilateral to the lesion (Figures 2, 3, 4B, 4C). These deviations from typically developing infants were consistently below the 95% confidence intervals of the regression lines. Notably, this increase was observed with age (between 9 and 19 weeks of corrected age), aligning with previous behavioral findings^7^, suggesting a progressive disruption of corticospinal development in the affected hemisphere.

As expected, the right arm (contralateral to the PVL lesion) in this infant was more affected, as indicated by both BabyOSCAR scores and the MPF/FA changes in the corresponding CST portions. Bilateral lower extremity involvement was also observed despite the unilateral lesion, potentially due to the early bilateral organization of lower extremity CST projections^43^. This pattern reinforces a direct link between periventricular injury and disruption of descending corticospinal pathways, highlighting the added value of early behavioral assessment. BabyOSCAR scores indicated bilateral lower extremity involvement, which may not have been fully anticipated from imaging alone. Together, these complementary measures support the use of pathway-specific imaging and motor assessment to anticipate the distribution of later motor involvement, thereby informing more timely and targeted support for children who show early signs of atypical motor development.

Further analysis of g-ratio, which reflects the ratio of inner axonal diameter to outer fiber diameter (an indicator of myelination efficiency), showed a larger developmental decrease in the PLIC compared to the corpus callosum in typically developing infants (Figure 5B), approaching optimal adult values around 0.7 ^39, 40^. In infants with CP, the g-ratio in the ipsilateral PLIC was consistently above the 95% confidence interval line at both 9 and 19 weeks corrected age, indicating suboptimal myelination efficiency in the affected tract. This deviation correlated with the increasing deviation in BabyOSCAR scores (Figure 5C). Conversely, the contralateral PLIC g-ratio remained within the typical range, further supporting the lesion-specific impact.

The MPF measure in the corpus callosum of the infant with CP was within the typical range, while FA in the corpus callosum was significantly lower (Figure 3). This suggests a dissociation in the impact of perinatal injury on different aspects of white matter maturation in commissural tracts compared to projection tracts, or potentially subtle compensatory mechanisms in myelination not fully captured by FA alone. Similarly, while cortical myelination generally increases with age (Figure 7A), the observed increase in cortical myelination contralateral to the lesion in the CP infant (Figure 7B, C) is a notable finding. This could represent a compensatory plastic response to the unilateral injury, where the uninjured hemisphere attempts to take on greater functional responsibility, potentially through enhanced myelination. Crucially, the increased myelination observed in the rubrospinal tract of the CP infant (Figure 6) is particularly noteworthy. This finding may reflect increased reliance on a compensatory pathway in response to CST injury. This mechanism has been observed in various neurological conditions ^34^, including increased reticulospinal excitability in adults with cerebral palsy^44^. These findings underscore the potential for alternate motor pathways, such as the rubrospinal or reticulospinal tracts, to support motor function when CST integrity is compromised. Additionally, these findings support the ongoing need to further evaluate and validate these measures in a population of young infants with CP

### Differentiating Early Motor Delay from CP

In contrast, one infant in our cohort who showed gross motor delay at 18 months (Bayley Scales >2 SD below the mean) but achieved independent walking by age two exhibited BabyOSCAR scores and CST imaging metrics (MPF, FA) within the 95% confidence intervals for typical development. This case suggests that early motor delay in the absence of structural CST abnormalities may reflect transient developmental differences rather than permanent injury. The ability to distinguish such cases from those with evolving CP through early imaging and motor assessments could enhance prognostic precision and guide more individualized intervention strategies.

### Limitations and Future Directions

The primary limitation of this study is the small sample size, particularly the inclusion of only one infant with cerebral palsy. While this single case provides compelling preliminary evidence, it limits the generalizability of our findings regarding CP. Future studies must incorporate larger cohorts of infants with various types and severities of perinatal brain injury to confirm these observations and establish the clinical utility of MPF and g-ratio as early biomarkers for motor impairment.

Longitudinal follow-up of these cohorts will be crucial to correlate early MRI findings with long-term motor and cognitive outcomes. Furthermore, investigating the specific mechanisms underlying the observed compensatory changes in the contralateral hemisphere and the rubrospinal tract, as well as the dissociation between MPF and FA in the corpus callosum, warrants further research. Incorporating multimodal imaging, and specifically advanced myelination development techniques such as MPF, holds significant promise for enhancing the prognostic accuracy of MRI for motor outcomes in infants with prematurity and for providing valuable insights in other developmental brain injury. A combination of these multimodal modalities, such as myelin imaging and fiber tract density estimation by advanced diffusion imaging, may further provide functional insight into the amount, efficiency, and timing of the development of motor control. By identifying structural markers that align with emerging selective motor control, this work supports the biological rationale for early identification and may help justify the timing and targeting of interventions in infants with atypical motor development.

## Data Availability

All data produced in the present study are available upon reasonable request to the authors

